# Electronic Health Record-Based Machine-Learning for Cardiac Arrhythmia Identification in Adults with High-Risk Chronic Disease: Model Development and Validation Study

**DOI:** 10.64898/2026.10.30.26366947

**Authors:** Mansi Goel, Shelby Matlock, Yasmin Karasu Benyes, Nanthini Narayanan, Jeremy Howard, Thomas George Brazeal, Colin Cogan, Eric Navarro, Andrei Petrus

## Abstract

**Background:** Cardiac arrhythmias may remain unrecognized in adults with chronic disease. Established risk scores primarily address atrial fibrillation.

**Objective:** To develop and internally evaluate an electronic health record (EHR)–based machine-learning model to identify patients more likely to receive a documented cardiac arrhythmia diagnosis.

**Methods:** We retrospectively evaluated EHR data (2018–2025) from one healthcare organization among adults with type 2 diabetes, chronic obstructive pulmonary disease, chronic kidney disease, or coronary artery disease. Logistic regression, random forest, and XGBoost were compared. XGBoost was selected using internal validation, reduced to 30 features using Shapley additive explanations, and evaluated in a holdout set with data ending one day before the index date. Sensitivity analyses excluded information recorded within 30, 90, or 180 days before the index date. An atrial fibrillation/flutter-specific analysis compared XGBoost with CHARGE-AF and HARMS_2_-AF.

**Results:** Among 30,356 patients, 6,835 (22.5%) had a qualifying arrhythmia diagnosis. In 3,051 holdout patients, XGBoost achieved an area under the receiver operating characteristic curve (AUROC) of 0.894 (95% CI 0.879–0.908) and an area under the precision-recall curve of 0.792 (95% CI 0.763–0.822). At a 0.5 threshold, sensitivity was 0.564, specificity 0.971, and precision 0.857. AUROC remained 0.896–0.897 across earlier cutoffs. In the common score-calculable atrial fibrillation/flutter subset (*n* = 2,436), AUROCs were 0.883 for XGBoost, 0.712 for CHARGE-AF, and 0.554 for HARMS_2_-AF.

**Conclusions:** XGBoost demonstrated strong internal discrimination for documented arrhythmias. External validation and prospective evaluation are needed to assess generalizability and clinical utility.

## 1. INTRODUCTION

Cardiac arrhythmias comprise a heterogeneous group of abnormalities in cardiac impulse formation and conduction, ranging from asymptomatic or intermittent rhythm disturbances to clinically consequential tachyarrhythmias and conduction disorders.[1, 2, 3] Because many arrhythmias are episodic or minimally symp-tomatic, diagnosis may occur incidentally or only after patients develop symptoms or related cardiovascular complications.[3]

The burden of arrhythmia is particularly relevant among patients with chronic cardiometabolic and cardiopulmonary disease. In a large study of patients with diabetes mellitus, atrial fibrillation was present in 14.9% and atrial flutter in 4.0%, compared with 10.3% and 2.5%, respectively, among controls.[4] In the Chronic Renal Insufficiency Cohort, atrial fibrillation was present in 18% of patients with chronic kidney disease (CKD) and in more than 25% of those aged 70 years or older.[5] Among patients with coronary artery disease (CAD), atrial fibrillation prevalence has been estimated at 0.2%–5%.[6] In a study of patients undergoing pulmonary function testing and Holter monitoring, those with chronic obstructive pulmonary disease (COPD) had higher frequencies of atrial fibrillation/atrial flutter (AF) (23.3% vs 11.0%), non-sustained ventricular tachycardia (13.0% vs 5.9%), and sustained ventricular tachycardia (1.6% vs 0.9%) than those without COPD.[7] These associations are biologically plausible: type 2 diabetes mellitus (T2DM) has been linked to autonomic, metabolic, inflammatory, and structural changes;[8] CKD to electrolyte and volume disturbances, autonomic dysfunction, and myocardial remodeling;[9] CAD to ischemia and myocardial scar;[1] and COPD to hypoxemia, inflammation, autonomic activation, and cardiac remodeling.[10] Together, T2DM, CKD, CAD, and COPD therefore define a clinically relevant population in whom arrhythmia risk is enriched and in whom scalable risk identification may be useful.

AF is the most common sustained cardiac arrhythmia and carries substantial risks of thromboembolic stroke, heart failure, and other cardiovascular morbidity.[11] However, AF represents only one component of the clinically relevant rhythm-disorder burden. Conduction disorders, paroxysmal tachycardias, and other cardiac arrhythmias may also prompt further rhythm assessment and management. In this context, risk identification should be distinguished from diagnosis: an electronic health record (EHR)-based model may identify patients with a greater likelihood of subsequently receiving an arrhythmia diagnosis, but confirmation still requires appropriate rhythm assessment and clinical interpretation.

Established clinical risk scores primarily address incident AF. Cohorts for Heart and Aging in Genomic Epidemiology (CHARGE-AF) estimates 5-year risk of incident AF using age, race, height, weight, systolic and diastolic blood pressure, smoking status, antihypertensive treatment, diabetes, prior myocardial infarction, and heart failure.[12] The original model achieved a C-statistic of 0.765, with C-statistics of 0.664 and 0.705 in two external validation cohorts.[12] HARMS_2_-AF is a weighted score based on hypertension, age, body mass index, male sex, sleep apnea, smoking, and alcohol consumption.[13] It achieved an area under the receiver operating characteristic curve (AUROC) of 0.782 in the UK Biobank and 0.757 in external validation in the Framingham Heart Study.[13] These scores provide clinically relevant benchmarks for AF risk prediction, but neither was developed for a broader outcome encompassing multiple cardiac arrhythmia categories or specifically for a high-risk population defined by T2DM, COPD, CKD, or CAD.

Several studies have investigated whether routinely collected EHR data and machine-learning (ML) methods can support arrhythmia risk prediction. Tiwari et al. evaluated more than 2.2 million EHR records for 6-month incident AF and compared logistic regression, random forest, neural networks, and gradient boosting; AUROCs ranged from 0.762 for gradient boosting to 0.806 for logistic regression, while the selected neural-network approach achieved an AUROC of 0.800 and provided only modest improvement over a model based on established AF risk factors.[14] Guo et al. used longitudinal EHR measures to predict a broader cardiac dysrhythmia outcome and found that a long short-term memory model achieved an AUROC of 0.76, compared with 0.71 for a deep neural network, 0.67 for random forest, and 0.66 for logistic regression.[15] Zhang et al. subsequently developed a statewide EHR model for 1-year incident cardiac dysrhythmia defined using International Classification of Diseases, Tenth Revision, Clinical Modification (ICD-10-CM) categories I47–I49. Among multiple architectures, prospective AUROCs were 0.827 for an ensemble model, 0.808 for eXtreme Gradient Boosting (XGBoost), 0.807 for a feed-forward neural network, and 0.695 for random forest.[16] For AF specifically, the FIND-AF random forest model achieved an AUROC of 0.824 for 6-month incident AF in UK primary-care EHR data[17] and was subsequently externally validated in more than two million patients in Israel with an AUROC of 0.871.[18] Taken together, these studies show that EHR-based arrhythmia prediction is feasible, but reported performance varies with the population, outcome definition, prediction horizon, and model architecture.

Prior studies have addressed both AF and broader dysrhythmia prediction,[14, 15, 16, 17, 18] but not in the combination evaluated here: a chronic-disease–enriched cohort, a broad I44–I49 outcome, longitudinal EHR features, and an AF-specific comparison with established clinical scores. The closest broad-outcome study evaluated I47–I49 in a general statewide population rather than a cohort defined by T2DM, COPD, CKD, or CAD.[16]

In this study, we retrospectively developed and compared logistic regression, random forest, and XGBoost models using EHR information recorded before the assigned index date among adults with at least one documented diagnosis of T2DM, COPD, CKD, or CAD. The primary outcome was the first recorded qualifying diagnosis within ICD-10-CM categories I44, Atrioventricular and left bundle-branch block; I47, Paroxysmal tachycardia; I48, Atrial fibrillation and flutter; or I49, Other cardiac arrhythmias, including all subcodes within each category.

### 1.1 Study Objectives

To develop and internally evaluate an EHR-based ML model to identify, among adults with a T2DM, COPD, CKD or CAD comorbidity, those patients with a greater likelihood of subsequently receiving a documented cardiac arrhythmia diagnosis in their available EHR data. We compared logistic regression, random forest, and XGBoost and evaluated the selected model on a held-out test set using records available up to one day before the index date, with sensitivity analyses excluding the final 30, 90, and 180 days. As a secondary analysis, XGBoost was compared with CHARGE-AF and HARMS_2_-AF after restricting the outcome to AF. [12, 13]

## 2. MATERIALS AND METHODS

### 2.1 Study design and data source

We conducted a retrospective cohort study to develop and internally validate an ML model for identifying patients at elevated risk for clinically documented cardiac arrhythmia. Documented arrhythmia, defined by ICD-10-CM diagnosis codes,[19] served as the prediction target and as a proxy for the presence of arrhythmia for supervised model training. The intended use is to identify patients without a documented qualifying arrhythmia who may warrant prioritization for further clinical evaluation. The model output is intended for risk stratification and should not be interpreted as establishing an arrhythmia diagnosis. All analyses were performed using EHR data from Desert Oasis Healthcare, covering the period 2018 through 2025.

### 2.2 Ethical Considerations

The machine-learning models were developed and the associated analyses were performed as part of a healthcare provider’s quality improvement activities under a business associate agreement. Subsequent interpretation and reporting of the existing results did not involve protected health information (PHI) or require access to source EHR records or patient-level datasets. This subsequent work involved no patient recruitment, direct patient contact, or changes to clinical care. The BRANY Institutional Review Board reviewed the study protocol and determined the study to be exempt under 45 CFR § 46.104(d)(4)(ii) (protocol LH-ARR-001; BRANY IRB file 26-12-716-2819). Individual informed consent was not required for this exempt secondary research.

### 2.3 Study population

Eligible patients were aged 18 years or older and had, before any qualifying arrhythmia diagnosis, at least one qualifying diagnosis of T2DM (E11), COPD (J40–J44), CKD (N18), or CAD (I20–I25), including all subcodes within these categories.

### 2.4 Outcome definition

The primary outcome was a documented qualifying cardiac arrhythmia diagnosis in the EHR within one of the following ICD-10-CM categories: I44 (atrioventricular and left bundle-branch block), I47 (paroxysmal tachycardia), I48 (AF), and I49 (other cardiac arrhythmias), including all subcodes within each category. Patients with one or more qualifying codes were classified as arrhythmia-positive, with the earliest qualifying diagnosis determining the primary arrhythmia category; patients meeting the comorbidity eligibility criterion but without any qualifying arrhythmia code were classified as arrhythmia-negative.

### 2.5 Observation window and model evaluation

The eligible cohort included 6,835 arrhythmia-positive cases and 23,521 arrhythmia-negative controls, or 3.44 controls per case. The available EHR observation period was defined using diagnosis, vital-sign, and laboratory records. For cases, the index date was the first recorded qualifying arrhythmia diagnosis.

Prior to model development, 10% of patients were randomly assigned to a final holdout test set that was excluded from all model training, tuning, and internal validation. The remaining 90% of patients were partitioned into training and internal validation sets using stratified random sampling to preserve arrhythmia prevalence. The training, internal validation, and holdout sets included 21,844, 5,461, and 3,051 patients, respectively, with similar arrhythmia prevalence across partitions (Supplementary Table S1).

Features were restricted to data before each patient’s cutoff, randomly set 1–180 days before the index date during development. Primary holdout evaluation used a 1-day cutoff, with 30-, 90-, and 180-day cutoffs for temporal sensitivity analyses.

### 2.6 Feature creation and preprocessing

Candidate predictors were derived from demographic, diagnostic, laboratory, and vital-sign/anthropometric EHR domains. Diagnostic features and corresponding ICD-10-CM codes are provided in Supplementary Table S2, laboratory features and Logical Observation Identifiers Names and Codes (LOINC) identifiers in Supplementary Table S3, and vital-sign and anthropometric features in Supplementary Table S4.[20] Race and ethnicity were harmonized into two categories for modeling: White, non-Hispanic and Other racial and ethnic groups. Patients were classified as White, non-Hispanic only when both criteria were recorded; all others were grouped as Other racial and ethnic groups. Laboratory, vital-sign, and anthropometric measurements were standardized and cleaned by harmonizing units according to Unified Code for Units of Measure (UCUM) conventions,[21] and excluding values outside prespecified biologically plausible ranges; detailed coding and range specifications are provided in Supplementary Tables S3 and S4.

For each variable, we calculated the median of all available measurements (_MEDIAN); a long-term normalized change, defined as the difference between the last and first values divided by elapsed days (_NORM_DELTA); an analogous normalized change restricted to the 547 days preceding the final measurement (_NORM_DELTA_18MO); and the mean and median of the three most recent measurements (_MEAN_LAST3 and _MEDIAN_LAST3). When only one measurement was available in the relevant window, normalized change was assigned a value of zero.

After feature creation and preprocessing, the initial modeling dataset included 128 candidate features before feature selection and model training.

Missing laboratory, vital sign, and anthropometric features were median-imputed using feature-specific medians estimated from the training subset and applied unchanged to the validation and holdout sets to prevent information leakage. Distributions and pre-imputation missingness of final XGBoost features are reported in Supplementary Table S5.

### 2.7 Model comparison, selection and hyperparameter tuning

Three supervised learning algorithms were evaluated: logistic regression,[22] random forest,[23] and gradient boosted decision trees implemented using XGBoost.[24] Logistic regression was included as an interpretable linear baseline, while random forest and XGBoost were evaluated to capture potential nonlinear relationships and interactions among clinical features.

The data used for model development was partitioned as described above. All models were trained and assessed within the same development framework, using performance on the internal validation set to guide model selection. Hyperparameter tuning was performed for logistic regression, random forest, and XGBoost using exhaustive grid search with 3-fold stratified cross-validation on the training set. AUROC was used as the optimization metric. The hyperparameter search space is provided in Supplementary Table S6.

For each model, the hyperparameter configuration with the highest mean cross-validated AUROC was selected. The three tuned models were then evaluated on the internal validation set to compare performance during model development. AUROC was used as the primary scoring metric, with additional consideration given to precision, specificity, and sensitivity. XGBoost showed the highest AUROC and area under the precision-recall curve (AUPRC) and was therefore selected for feature reduction and final holdout evaluation.

### 2.8 SHAP-based feature selection and final model training

After hyperparameter tuning, SHapley Additive exPlanations (SHAP) values[25] were calculated across patients in the training data to estimate each feature’s contribution to the XGBoost model’s predictions. Features were ranked according to mean absolute SHAP value in the training data. The 30 highest-ranked features were retained to produce a more parsimonious final model and facilitate interpretation. The choice of 30 features was pragmatic rather than derived from a formal feature-count optimization procedure.

The XGBoost model with the top 30 features and the selected hyperparameter configuration was then trained on the combined training and internal validation sets, representing 90% of the full study cohort.

### 2.9 Final model evaluation

The final XGBoost model was evaluated on the prespecified holdout set using a 0.5 probability threshold. Performance was assessed using AUROC, AUPRC, sensitivity, specificity, precision, negative predictive value (NPV), F1 score, accuracy, and Brier score. The 30-, 90-, and 180-day cutoffs were evaluated as temporal sensitivity analyses. Final model performance was also examined separately across the four arrhythmia categories defined by the earliest qualifying ICD-10-CM diagnosis within I44, I47, I48, or I49, including all subcodes.

### 2.10 Model interpretability

Model interpretability was assessed using SHAP values for the final XGBoost model. For the 30 retained features, SHAP beeswarm plots and mean absolute SHAP importance rankings were generated to visualize feature contributions. These plots were used to examine the direction and magnitude of individual feature contributions to the model output. SHAP values were interpreted as model attributions and not as causal effects or estimates of absolute clinical risk.

### 2.11 Subgroup and fairness analyses

Subgroup analyses were conducted to assess whether final model performance was consistent across clinically and demographically relevant groups. Subgroups included recorded gender, race/ethnicity, and age. Recorded gender was evaluated as female and male, and race/ethnicity was evaluated using the two-level race/ethnicity feature. Age-based subgroup performance was evaluated using four categories: 18–39, 40–64, 65–79, and ≥80 years.

Fairness was evaluated using demographic parity difference and equalized odds difference, implemented with the Fairlearn library.[26] These metrics were used to assess between-group differences in positive prediction rates and error-rate patterns, respectively. Values closer to zero indicate more similar model behavior across groups.

### 2.12 Comparison with CHARGE-AF and HARMS**_2_**-AF

CHARGE-AF and HARMS_2_-AF were calculated as secondary clinical benchmarks for the AF component of the study outcome. The published CHARGE-AF equation was used to generate a 5-year absolute risk estimate for AF, whereas HARMS_2_-AF was evaluated as an integer point-based risk score. Because both scores were developed for AF prediction, this comparison was restricted to patients with diagnoses within I48, including all subcodes, as positive cases and patients from the primary arrhythmia-negative cohort as controls; patients whose qualifying outcome was a non-I48 arrhythmia were excluded.

All three approaches were evaluated in the same subset of holdout patients for whom both clinical scores were calculable. Discrimination was assessed using AUROC and AUPRC. Threshold-dependent performance was additionally reported at a probability threshold of 0.5 for XGBoost, CHARGE-AF 5-year risk thresholds of 2.5% and 5%,[27] and a HARMS_2_-AF threshold of ≥5 points.[13]

The common score-calculable comparison cohort was additionally characterized relative to the full holdout population with respect to demographic characteristics and outcome composition (Supplementary Table S7).

## 3. RESULTS

### 3.1 Cohort Characteristics

The patient selection process is summarized in Figure 1. The final cohort included 30,356 patients, of whom 6,835 (22.5%) were arrhythmia-positive and 23,521 (77.5%) were arrhythmia-negative. By primary arrhythmia category, 700 patients were classified under I44, 1542 under I47, 3284 under I48, and 1309 under I49. Demographic characteristics and qualifying comorbidities of the final study cohort are summarized in Table 1.

**Figure 1.**
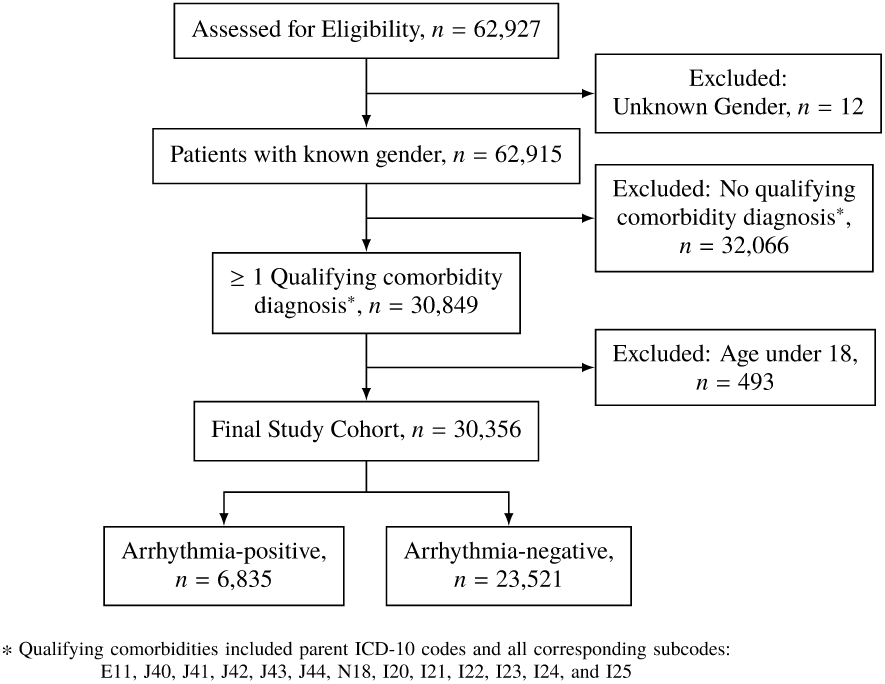
Cohort selection flow diagram. ICD-10: International Classification of Diseases, Tenth Revision.

**Table 1.** Demographic and qualifying comorbidity characteristics of the final study cohort by arrhythmia status. Continuous variables are reported as median (IQR), and categorical variables are reported as n (%). Percentages are calculated within each column. Qualifying comorbidities were not mutually exclusive; patients could have more than one qualifying condition. Patients were classified as White, non-Hispanic only if race was recorded as White and ethnicity was recorded as non-Hispanic; all other patients were grouped as Other racial and ethnic groups. IQR, interquartile range.

| **Characteristic** | **Overall cohort (n = 30,356)** | **Arrhythmia-negative (n = 23,521)** | **Arrhythmia-positive (n = 6,835)** |
|---|---|---|---|
| Age, median (IQR), years | 72.0 (64.0–79.0) | 70.0 (60.0–78.0) | 78.0 (72.0–84.0) |
| Age 18–39, n (%) | 1,778 (5.9) | 1,742 (7.4) | 36 (0.5) |
| Age 40–64, n (%) | 6,217 (20.5) | 5,700 (24.2) | 517 (7.6) |
| Age 65–79, n (%) | 14,809 (48.8) | 11,560 (49.1) | 3,249 (47.5) |
| Age ≥80, n (%) | 7,552 (24.9) | 4,519 (19.2) | 3,033 (44.4) |
| Female, n (%) | 15,474 (51.0) | 12,340 (52.5) | 3,134 (45.9) |
| Male, n (%) | 14,882 (49.0) | 11,181 (47.5) | 3,701 (54.1) |
| White, non-Hispanic, n (%) | 10,831 (35.7) | 7,494 (31.9) | 3,337 (48.8) |
| Other racial and ethnic groups, n (%) | 19,525 (64.3) | 16,027 (68.1) | 3,498 (51.2) |
| Type 2 diabetes mellitus, n (%) | 17,158 (56.5) | 13,671 (58.1) | 3,487 (51.0) |
| Chronic obstructive pul-monary disease, n (%) | 18,724 (61.7) | 14,364 (61.1) | 4,360 (63.8) |
| Chronic kidney disease, n (%) | 11,331 (37.3) | 7,433 (31.6) | 3,898 (57.0) |
| Coronary artery disease, n (%) | 12,876 (42.4) | 8,999 (38.3) | 3,877 (56.7) |

### 3.2 Model comparison and selection

XGBoost had the highest internal-validation AUROC (0.8954; 95% confidence interval (CI), 0.8843–0.9065) and AUPRC (0.7984; 95% CI, 0.7786–0.8186), followed by random forest and logistic regression (Table 2).

**Table 2.** Internal validation performance of candidate models. AUROC and AUPRC are threshold-independent metrics. Sensitivity, specificity, precision, negative predictive value, and F1 score were calculated at the default probability threshold of 0.5. All performance metrics are reported with 95% CI. AUROC, area under the receiver operating characteristic curve; AUPRC, area under the precision-recall curve; NPV, negative predictive value.

| **Metric** | **Logistic Regression** | **Random Forest** | **XGBoost** |
|---|---|---|---|
| AUROC (95% CI) | 0.8349 (0.8217–0.8481) | 0.8804 (0.8683–0.8926) | 0.8954 (0.8843–0.9065) |
| AUPRC (95% CI) | 0.6478 (0.6199–0.6770) | 0.7775 (0.7572–0.7982) | 0.7984 (0.7786–0.8186) |
| Sensitivity (95% CI) | 0.7363 (0.7109–0.7602) | 0.6400 (0.6127–0.6664) | 0.6424 (0.6152–0.6688) |
| Specificity (95% CI) | 0.7729 (0.7600–0.7853) | 0.9412 (0.9337–0.9479) | 0.9551 (0.9485–0.9610) |
| Precision (95% CI) | 0.4839 (0.4613–0.5066) | 0.7590 (0.7319–0.7840) | 0.8055 (0.7795–0.8291) |
| NPV (95% CI) | 0.9102 (0.9004–0.9191) | 0.9004 (0.8912–0.9089) | 0.9023 (0.8933–0.9107) |
| F1 score (95% CI) | 0.5840 (0.5625–0.6043) | 0.6944 (0.6717–0.7161) | 0.7148 (0.6931–0.7365) |

Comparative AUROC and AUPRC curves are shown in Figures 2 and 3, respectively. At the default 0.5 threshold, XGBoost also had the highest specificity, precision, and F1 score, whereas logistic regression had the highest sensitivity. XGBoost was therefore selected for feature reduction and final holdout evaluation.

**Figure 2.**
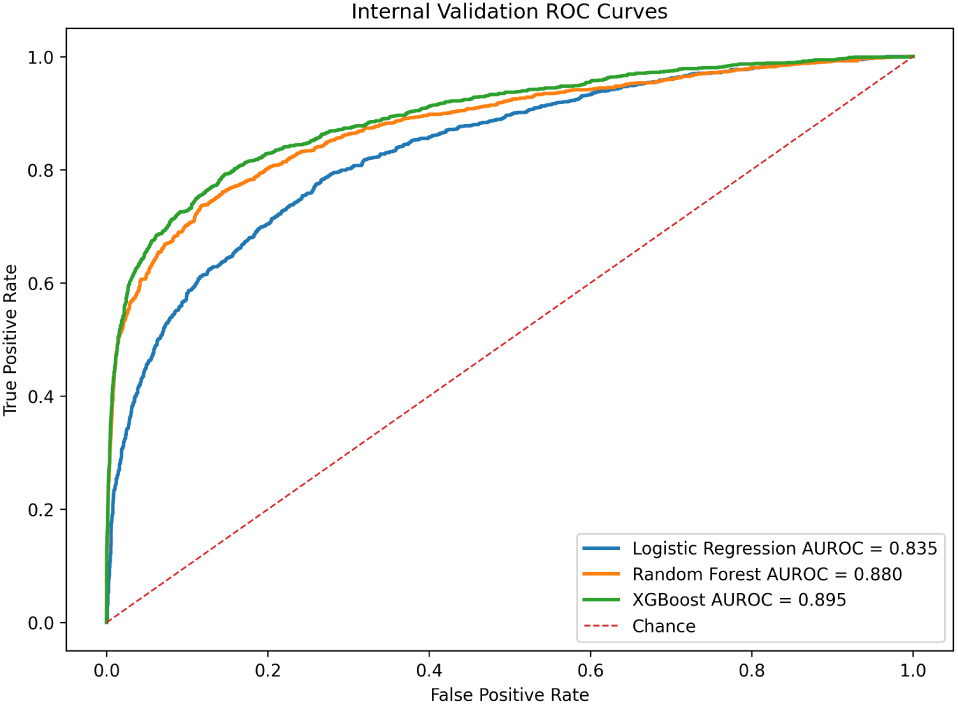
Receiver operating characteristic curves for logistic regression, random forest, and XGBoost models on the internal validation set. AUROC: area under the receiver operating characteristic curve.

**Figure 3.**
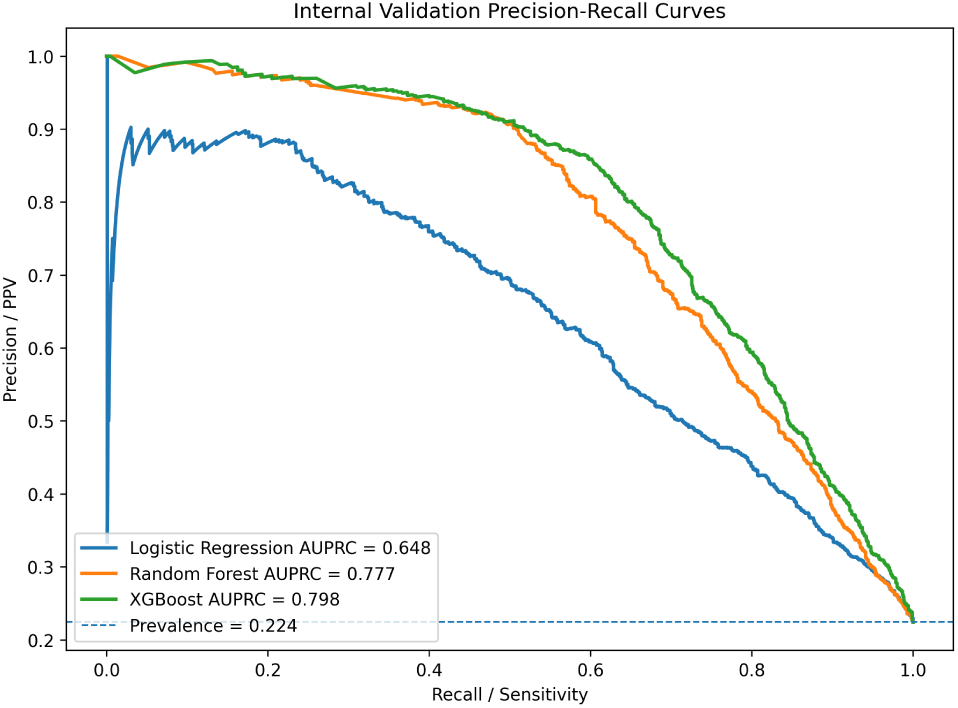
Precision-recall curves for logistic regression, random forest, and XGBoost models on the internal validation set. AUPRC: area under the precision-recall curve.

### 3.3 Final Model Evaluation

The final XGBoost model was trained using the top 30 SHAP-selected features and the selected hyperparameter configuration on the combined training and internal validation sets, then evaluated on the primary held-out test set with a 1-day pre-index cutoff. Performance metrics are summarized in Table 3. Holdout AUROC and AUPRC are shown in Figures 4 and 5, with the corresponding confusion matrix and calibration curve provided in Supplementary Figure S1 and Supplementary Figure S2.

**Figure 4.**
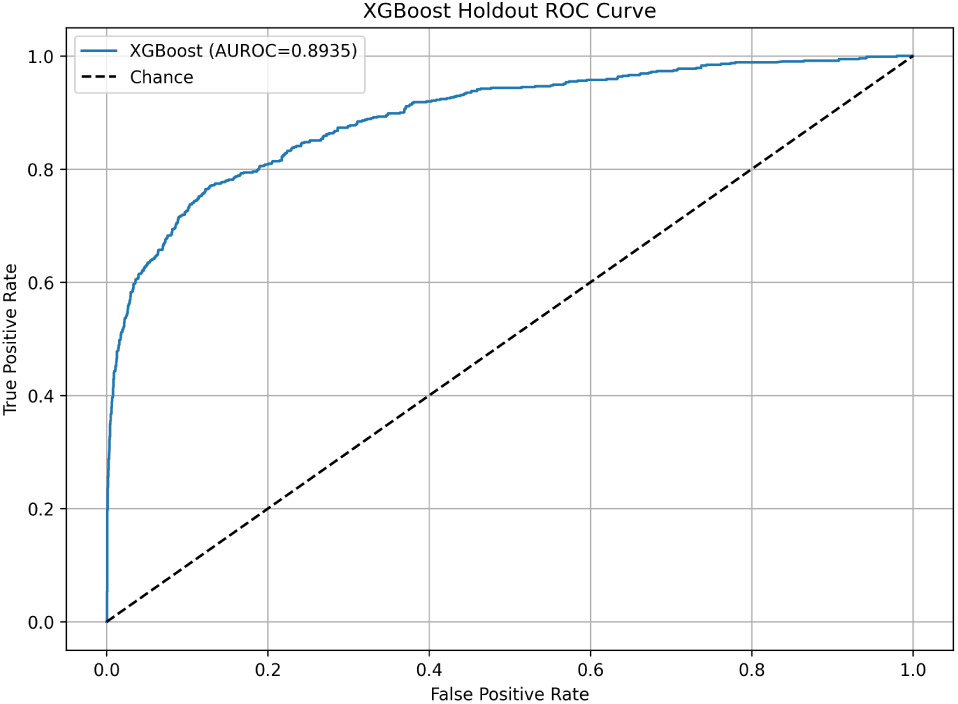
Receiver operating characteristic curve for the final XGBoost model on the holdout test set. AUROC: area under the receiver operating characteristic curve.

**Figure 5.**
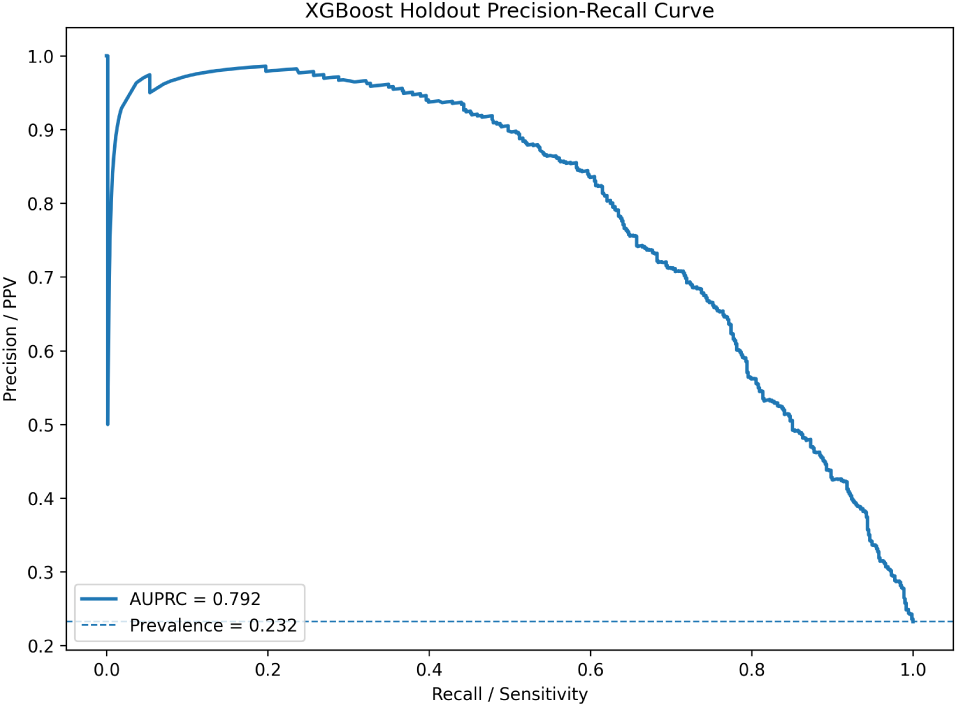
Precision-recall curve for the final XGBoost model on the holdout test set. AUPRC: area under the precision-recall curve; PPV: positive predictive value.

**Table 3.**
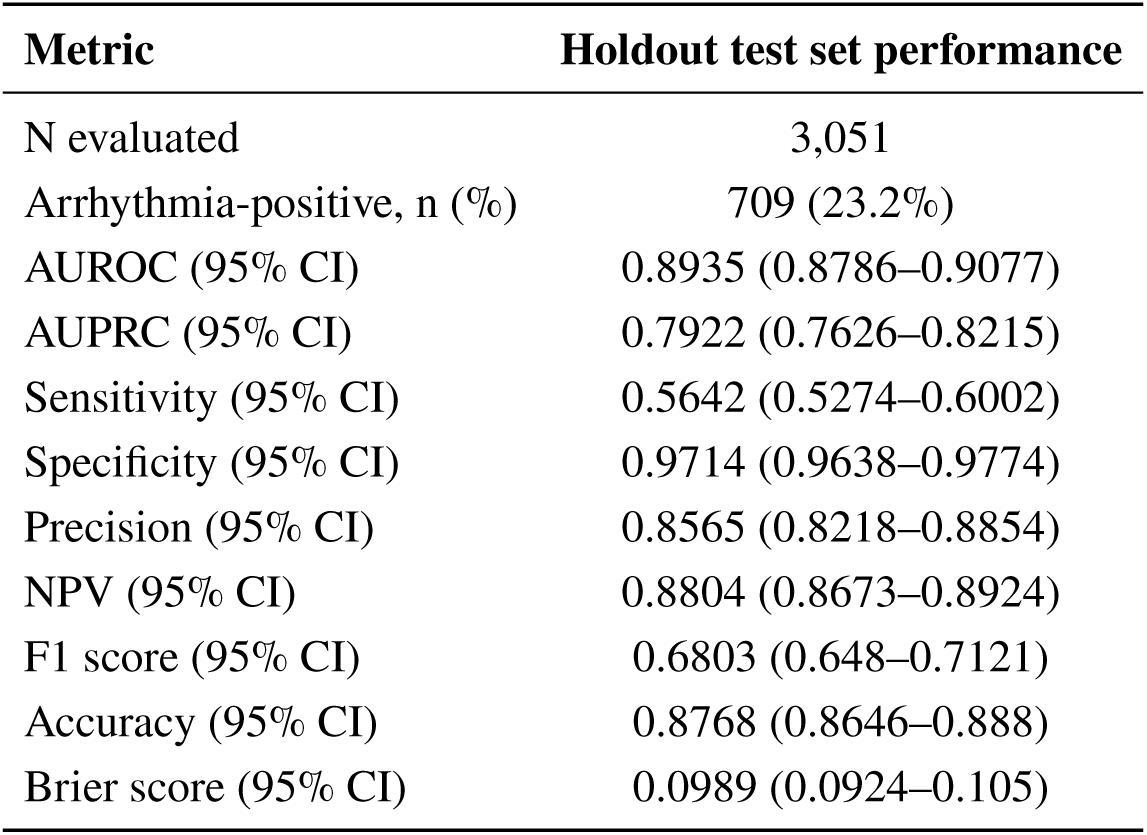
Final XGBoost model performance on the primary held-out test set. AUROC, AUPRC, and Brier score are threshold-independent metrics. Sensitivity, specificity, precision, negative predictive value, F1 score, and accuracy were calculated at the default probability threshold of 0.5. All performance metrics are reported with 95% CI. AUROC, area under the receiver operating characteristic curve; AUPRC, area under the precision-recall curve; NPV, negative predictive value.

All evaluations using earlier feature cutoffs included the same 3,051 holdout patients, including 709 (23.2%) arrhythmia-positive patients. Discrimination remained stable across temporal cutoffs. AUROC was 0.8935 at 1 day, 0.8968 at 30 days, 0.8963 at 90 days, and 0.8963 at 180 days. Full performance metrics are presented in Supplementary Table S8.

Performance across individual arrhythmia target groups is reported in Supplementary Table S9.

### 3.4 Final XGBoost Model SHAP Feature Importance and Model Explainability

SHAP-based feature importance was used to identify the predictors contributing most strongly to the final XGBoost model. Age had the highest mean absolute SHAP value and therefore the greatest influence on model predictions. Other influential features included demographic characteristics and laboratory-derived predictors, particularly hematologic and metabolic markers. Several top-ranked predictors represented normalized longitudinal changes in laboratory measurements rather than static values. Figure 6 presents the top 10 SHAP-ranked features, and Supplementary Figure S3 presents the complete set of 30 SHAP-selected features.

**Figure 6.**
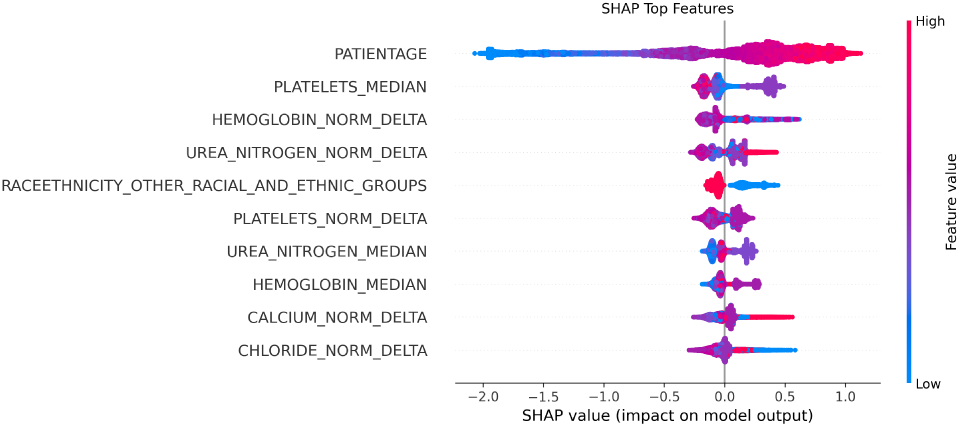
Shapley additive explanations beeswarm plot for the top 10 predictors in the final XGBoost model. Each point represents 1 patient; horizontal position indicates the direction and magnitude of that feature’s contribution to the model output, and the color scale encodes the underlying feature value from low to high.

### 3.5 Subgroup and fairness analyses

At the 0.5 classification threshold, demographic parity and equalized odds differences were 0.019 and 0.011 for gender and 0.115 and 0.129 for race/ethnicity, respectively. Subgroup performance results are presented in Supplementary Tables S10 and S11.

### 3.6 Comparison with CHARGE-AF and HARMS**_2_**-AF

The final XGBoost model was compared with CHARGE-AF and HARMS_2_-AF among holdout patients with sufficient data for score calculation. Their common calculable subset included 2,725 patients. Patients positive for non-I48 arrhythmias were excluded, resulting in a final AF-specific benchmark cohort of 2,436 patients, including 208 I48-positive patients (8.5%) and 2,228 arrhythmia-negative patients.

On the common AF-specific cohort, XGBoost demonstrated greater discrimination than CHARGE-AF and HARMS_2_-AF. Detailed comparative performance across the evaluated thresholds is presented in Table 4.

**Table 4.**
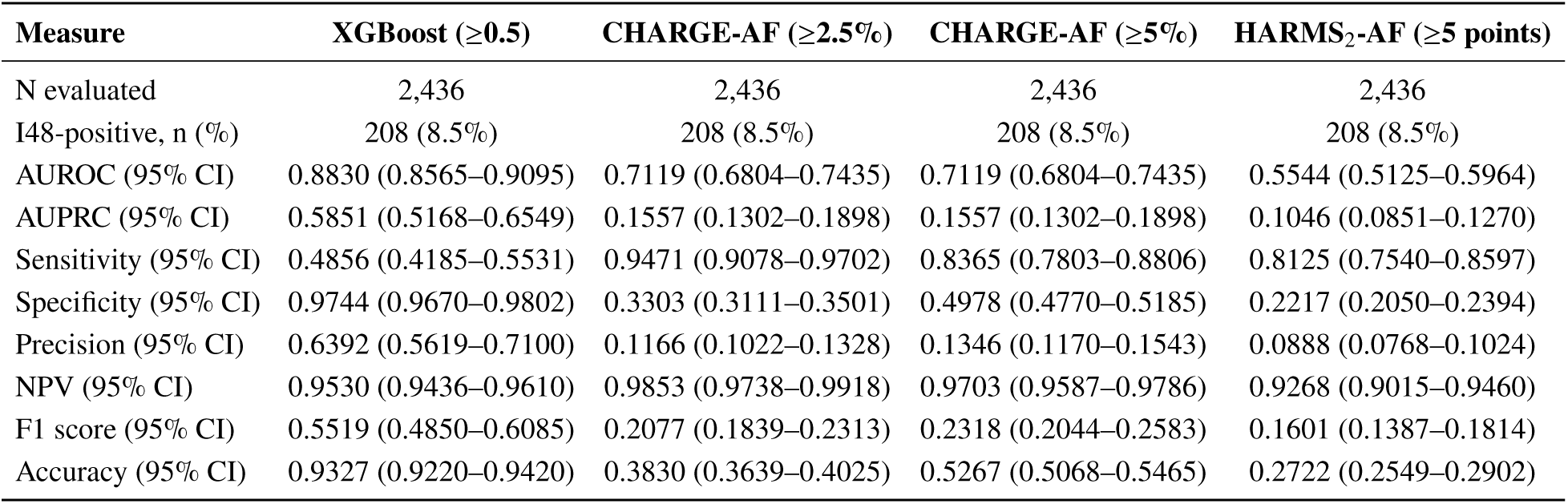
Comparison of XGBoost, CHARGE-AF, and HARMS_2_-AF for AF prediction in the final AF-specific benchmark cohort. Performance was evaluated on the same subset of held-out test patients for whom both CHARGE-AF and HARMS_2_-AF were calculable. For this comparison, analyses were restricted to AF (I48), including all subcodes as described in the Methods. Threshold-dependent metrics were calculated using a probability threshold of 0.5 for XGBoost, 5-year predicted AF risk thresholds of 2.5% and 5% for CHARGE-AF, and a score threshold of ≥5 points for HARMS_2_-AF. Brier score was not reported for HARMS_2_-AF because its raw point-based score does not represent a calibrated probability estimate. All performance metrics are reported with 95% CI. AF, atrial fibrillation; CHARGE-AF, Cohorts for Heart and Aging Research in Genomic Epidemiology; AUROC, area under the receiver operating characteristic curve; AUPRC, area under the precision-recall curve; NPV, negative predictive value.

| **Measure** | **XGBoost (≥0.5)** | **CHARGE-AF (≥2.5%)** | **CHARGE-AF (≥5%)** | **HARMS<sub>2</sub>-AF (≥5 points)** |
|---|---|---|---|---|
| N evaluated | 2,436 | 2,436 | 2,436 | 2,436 |
| I48-positive, n (%) | 208 (8.5%) | 208 (8.5%) | 208 (8.5%) | 208 (8.5%) |
| AUROC (95% CI) | 0.8830 (0.8565–0.9095) | 0.7119 (0.6804–0.7435) | 0.7119 (0.6804–0.7435) | 0.5544 (0.5125–0.5964) |
| AUPRC (95% CI) | 0.5851 (0.5168–0.6549) | 0.1557 (0.1302–0.1898) | 0.1557 (0.1302–0.1898) | 0.1046 (0.0851–0.1270) |
| Sensitivity (95% CI) | 0.4856 (0.4185–0.5531) | 0.9471 (0.9078–0.9702) | 0.8365 (0.7803–0.8806) | 0.8125 (0.7540–0.8597) |
| Specificity (95% CI) | 0.9744 (0.9670–0.9802) | 0.3303 (0.3111–0.3501) | 0.4978 (0.4770–0.5185) | 0.2217 (0.2050–0.2394) |
| Precision (95% CI) | 0.6392 (0.5619–0.7100) | 0.1166 (0.1022–0.1328) | 0.1346 (0.1170–0.1543) | 0.0888 (0.0768–0.1024) |
| NPV (95% CI) | 0.9530 (0.9436–0.9610) | 0.9853 (0.9738–0.9918) | 0.9703 (0.9587–0.9786) | 0.9268 (0.9015–0.9460) |
| F1 score (95% CI) | 0.5519 (0.4850–0.6085) | 0.2077 (0.1839–0.2313) | 0.2318 (0.2044–0.2583) | 0.1601 (0.1387–0.1814) |
| Accuracy (95% CI) | 0.9327 (0.9220–0.9420) | 0.3830 (0.3639–0.4025) | 0.5267 (0.5068–0.5465) | 0.2722 (0.2549–0.2902) |

## 4. DISCUSSION

In this retrospective EHR-based cohort study of adults with T2DM, COPD, CKD, or CAD, we developed and internally evaluated a ML model for identifying patients at elevated risk of clinically documented cardiac arrhythmia. The final XGBoost model used 30 SHAP-selected features spanning demographic, diagnostic, laboratory, vital sign, and anthropometric domains and demonstrated strong discrimination in the primary held-out test set. Model discrimination was similar across the 30-, 90-, and 180-day feature cutoffs in the holdout set.

Among the three candidate models, XGBoost had the highest discrimination. Its AUROC was also numerically higher than that reported in several prior EHR-based studies of AF and broader cardiac dysrhythmia prediction,[14, 15, 16, 17, 18] although cross-study comparisons should be interpreted cautiously given differences in study populations, outcome definitions, prediction horizons, feature sets, and validation designs. The principal distinction of the present study is its evaluation of a broader documented-arrhythmia outcome among adults with T2DM, COPD, CKD, or CAD, using longitudinal EHR information. It additionally provides an AF-specific head-to-head comparison with established clinical risk scores in a common calculable subset.

Within the AF-specific comparison, XGBoost demonstrated greater discrimination than CHARGE-AF and HARMS_2_-AF. CHARGE-AF and HARMS_2_-AF use a limited set of predefined clinical risk factors, whereas the XGBoost model incorporated a broader set of EHR-derived features, including longitudinal laboratory and vital-sign summaries. CHARGE-AF was developed to estimate incident AF risk over a 5-year horizon, and HARMS_2_-AF was developed and externally validated for new-onset AF in community-based populations.[12, 13] XGBoost showed greater relative discrimination within this comorbidity-enriched cohort and serves as a secondary cohort-specific benchmark. The EHR model generated predictions for the full holdout cohort, whereas the clinical score comparison was limited to patients with the required input variables. This distinction is relevant when applying prediction approaches to routinely collected EHR data, in which missingness and patterns of data collection are common methodological considerations.[28, 29]

SHAP analysis provided insight into model predictions. Age was the strongest contributor, consistent with the previously established increase in arrhythmia incidence, particularly AF, with advancing age.[30] Multiple hematologic and renal/metabolic measures, including platelet, hemoglobin, blood urea nitrogen, calcium, and chloride, were also influential. Several highly ranked predictors represented time-standardized changes in laboratory measurements, indicating that the fitted model used longitudinal information in addition to static summaries. SHAP does not, however, quantify the independent predictive value of longitudinal features.

Model performance was generally similar across recorded gender and race/ethnicity groups. At the default threshold, between-group differences were smaller for recorded gender than for race/ethnicity. The latter finding may reflect variation in outcome prevalence and healthcare utilization,[28, 29] which can influence feature availability and outcome documentation in EHRs. The two-level race/ethnicity classification also prevented more granular assessment.

Age-stratified performance was less stable in younger adults: only five positive cases were available among patients aged 18–39 years. Performance in this subgroup therefore remains uncertain.

At the 0.5 threshold, the model traded sensitivity for high specificity and precision, yielding a relatively small, enriched subgroup that could potentially be prioritized for further clinical evaluation. The appropriate operating threshold would depend on the intended clinical workflow and the relative consequences of false-positive and false-negative predictions. ICD-10-CM diagnoses served as the study outcome, and confirmation of an arrhythmia requires appropriate rhythm assessment and clinical interpretation. Future prospective studies could evaluate model-guided rhythm assessment for earlier identification of clinically confirmed arrhythmia.

Together, these findings support further evaluation of longitudinal EHR-based ML risk identification for prioritizing clinical assessment in populations with cardiometabolic and cardiopulmonary disease burden.

### 4.1 Limitations

This study has several limitations. First, model development and evaluation were performed using data from a single healthcare organization. A prespecified holdout set provided independent internal evaluation, but external validation is needed to assess transportability across populations, care settings, coding practices, and EHR data availability.

Second, the outcome was based on ICD-10-CM diagnosis codes rather than adjudicated electrocardiographic or rhythm-monitoring findings.[31] The first qualifying code may not represent biological disease onset, and some patients classified as negative may have had undocumented arrhythmias. Positive labels may also reflect historical, miscoded, or unconfirmed diagnosis. The results therefore represent prediction of documented arrhythmia rather than adjudicated incident arrhythmia.

Third, the study did not estimate absolute arrhythmia risk over a fixed follow-up interval from a common prediction time. The temporal sensitivity analyses assessed discrimination at earlier feature cutoffs. Differences in observation history or healthcare utilization may still have influenced performance.

Fourth, routinely collected EHR data are subject to informative presence and missingness.[28, 29, 32] Patients with greater healthcare utilization generate more measurements and have more opportunities for an arrhythmia diagnosis to be recorded. Median imputation enabled complete scoring but did not capture informative missingness, while longitudinal summaries may reflect differences in measurement density.

Finally, the reported 0.5 probability threshold was not optimized for clinical utility. Future external and prospective evaluation could assess discrimination, calibration, workflow-specific thresholds, and clinical utility including decision-curve analysis.[33]

## 5. CONCLUSION

An EHR-based XGBoost model using routinely collected longitudinal clinical information demonstrated strong internal discrimination for documented cardiac arrhythmias in adults with T2DM, COPD, CKD, or CAD. Discrimination remained similar after progressively excluding more proximal EHR information, and the model showed greater discrimination than CHARGE-AF and HARMS_2_-AF in a secondary AF-specific benchmark. These findings support further evaluation of EHR-based arrhythmia risk identification.

## INSTITUTIONAL REVIEW BOARD STATEMENT

The BRANY Institutional Review Board reviewed the study protocol and determined that it qualified as exempt secondary research under 45 CFR § 46.104(d)(4)(ii) (protocol LH-ARR-001; BRANY IRB file 26-12-716-2819). The reviewed work involved the interpretation and reporting of existing non-identifiable results, from which individuals’ identities could not readily be ascertained directly or through linked identifiers. No individuals were recruited or contacted, and no re-identification was undertaken to produce the present study. The work did not involve protected health information or require access to source EHR records.

## INFORMED CONSENT STATEMENT

Individual informed consent was not required for this exempt secondary research under 45 CFR § 46.104(d)(4)(ii). The reviewed activity involved no patient recruitment, direct patient contact, or re-identification. No identifiable individual information is reported in this manuscript.

## DATA AVAILABILITY STATEMENT

The patient-level EHR data analyzed in this study are not publicly available because of patient privacy requirements, data-use agreements, and proprietary restrictions. The data were obtained by Lucem Health from Desert Oasis Healthcare under a pilot program evaluating the Reveal for Arrhythmia solution in the Desert Oasis Healthcare patient population. Under the applicable data-use restrictions, the dataset cannot be made available to external researchers.

## AUTHOR CONTRIBUTIONS

**Mansi Goel:** Data curation, Formal analysis, Investigation, Methodology, Software, Validation, Visualization, Writing – original draft, Writing – review & editing (lead).

**Shelby Matlock:** Data curation, Formal analysis, Investigation, Methodology, Software, Validation, Writing – original draft, Writing – review & editing (supporting).

**Yasmin Karasu Benyes:** Data curation, Formal analysis, Investigation, Methodology, Software, Validation, Writing – review & editing (supporting).

**Nanthini Narayanan:** Data curation, Formal analysis, Investigation, Methodology, Software, Validation, Writing – review & editing (supporting).

**Jeremy Howard:** Data curation, Software, Writing – review & editing (supporting).

**Thomas George Brazeal:** Data curation, Writing – review & editing (supporting).

**Colin Cogan:** Project administration, Resources, Writing – review & editing (supporting).

**Eric Navarro:** Funding acquisition, Resources, Writing – review & editing (supporting).

**Andrei Petrus:** Conceptualization, Supervision, Writing – review & editing (supporting).

## ABBREVIATIONS

AF: atrial fibrillation/flutter
AUPRC: area under the precision-recall curve
AUROC: area under the receiver operating characteristic curve
CAD: coronary artery disease
CHARGE-AF: Cohorts for Heart and Aging Research in Genomic Epidemiology
CKD: chronic kidney disease
COPD: chronic obstructive pulmonary disease
EHR: electronic health record
HARMS_2_-AF: hypertension, age, raised body mass index, male sex, sleep apnea, smoking, and alcohol
ICD-10-CM: International Classification of Diseases, Tenth Revision, Clinical Modification
ML: machine learning
NPV: negative predictive value
SHAP: SHapley Additive exPlanations
T2DM: type 2 diabetes mellitus
XGBoost: eXtreme Gradient Boosting

## SUPPLEMENTARY MATERIALS

**Supplementary Table S1.**
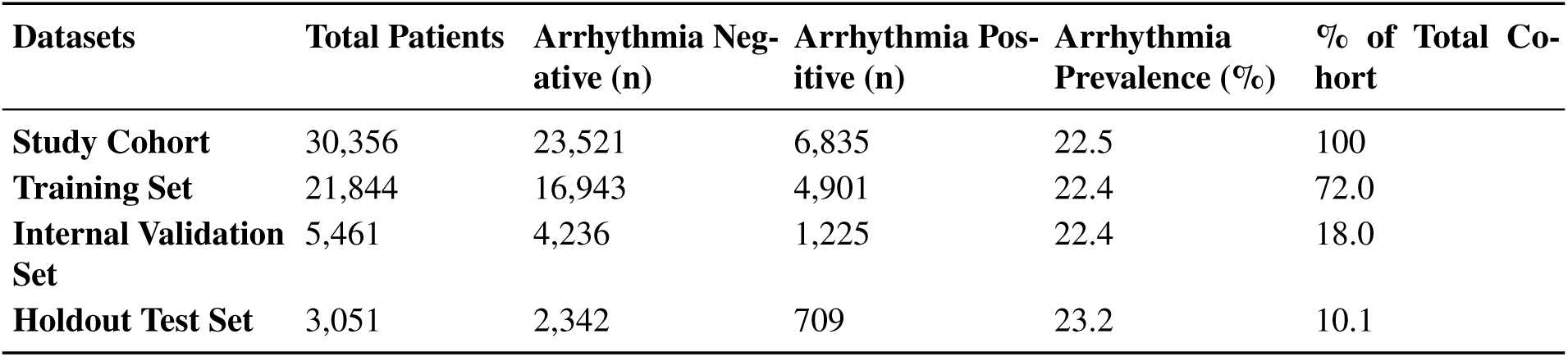
Dataset partitioning and arrhythmia prevalence across analytic splits. Percentages for arrhythmia prevalence are calculated within each dataset split. Percentages of total cohort are calculated using the full study cohort denominator of 30,356 patients.

| **Datasets** | **Total Patients** | **Arrhythmia Neg-ative (n)** | **Arrhythmia Pos-itive (n)** | **Arrhythmia Prevalence (%)** | **% of Total Co-hort** |
|---|---|---|---|---|---|
| **Study Cohort** | 30,356 | 23,521 | 6,835 | 22.5 | 100 |
| **Training Set** | 21,844 | 16,943 | 4,901 | 22.4 | 72.0 |
| **Internal Validation Set** | 5,461 | 4,236 | 1,225 | 22.4 | 18.0 |
| **Holdout Test Set** | 3,051 | 2,342 | 709 | 23.2 | 10.1 |

**Supplementary Table S2.** ICD-10-CM definitions for diagnosis-derived comorbidities and clinical history features. Listed ICD-10-CM codes, prefixes, and ranges were matched using prefix-based logic and include all subcodes unless an exact code is specified. T2DM, COPD, CKD, and CAD were used to define cohort eligibility and were not included as model predictors. ICD-10-CM, International Classification of Diseases, Tenth Revision, Clinical Modification; T2DM, type 2 diabetes mellitus; COPD, chronic obstructive pulmonary disease; CKD, chronic kidney disease; CAD, coronary artery disease.

| **Comorbidity or clinical feature** | **Identifying ICD-10-CM codes/prefixes** |
|---|---|
| **Anemia** | D50–D64 |
| **Cancer** | C00–D49 |
| **Cardiac surgery** | Z95.2, Z48.812 |
| **Colorectal surgery** | Z48.815 |
| **Prior COVID-19 diagnosis** | U07.1, U07.2, U09.9, U49, J12.81, J12.82, Z11.52, Z20.822, Z86.16, Z28.31, Z28.310, Z28.311, B97.29 |
| **Deep venous thrombosis** | I82 |
| **Dyslipidemia** | E78 |
| **Electrophysiology procedure, atria** | Z45.01, Z45.02 |
| **Heart failure** | I50 |
| **Hypertension** | I10, I11, I12, I13, I15, I1A |
| **Hyperthyroidism** | E05 |
| **Hypertrophic cardiomyopathy** | I42.1, I42.2 |
| **Infective endocarditis** | I33 |
| **Infection** | A04, J10, J15, A41 |
| **Inflammatory bowel disease** | K50, K51 |
| **Chronic ischemic heart disease** | I25 |
| **Mitral regurgitation** | I34.0 |
| **Mitral stenosis** | I05.0 |
| **Myocarditis** | I40, I41 |
| **Multiple valve disease** | I08 |
| **Pericarditis** | I30, I32 |
| **Pulmonary embolus** | I26 |
| **Pulmonary hypertension** | I27 |
| **Stroke or transient ischemic attack** | I63, I64, G45 |
| **Respiratory surgery** | Z48.813 |
| **Tobacco use** | Z72.0, Z77.22, Z87.891, F17 |
| **Alcohol abuse** | F10 |
| **Drug abuse** | F11, F12, F13, F14, F15, F16, F18, F19 |
| **Valvular heart disease** | I05, I09, I34, I39 |
| **Vascular dementia** | F01 |
| **Systolic heart failure** | I50.2 |
| **Congestive heart failure** | I50.2, I50.3, I50.4 |
| **Transient ischemic attack** | G45.8, G45.9 |
| **Stroke** | I63 |
| **Thromboembolism** | I74 |
| **Myocardial infarction** | I21, I22, I23 |
| **Peripheral artery disease** | I73 |
| **Atherosclerosis** | I70 |
| **Chronic obstructive pulmonary disease** | J40–J44 |
| **Type 2 diabetes mellitus** | E11 |
| **Chronic kidney disease** | N18 |
| **Coronary artery disease** | I20, I21, I22, I23, I24, I25 |

**Supplementary Table S3.**
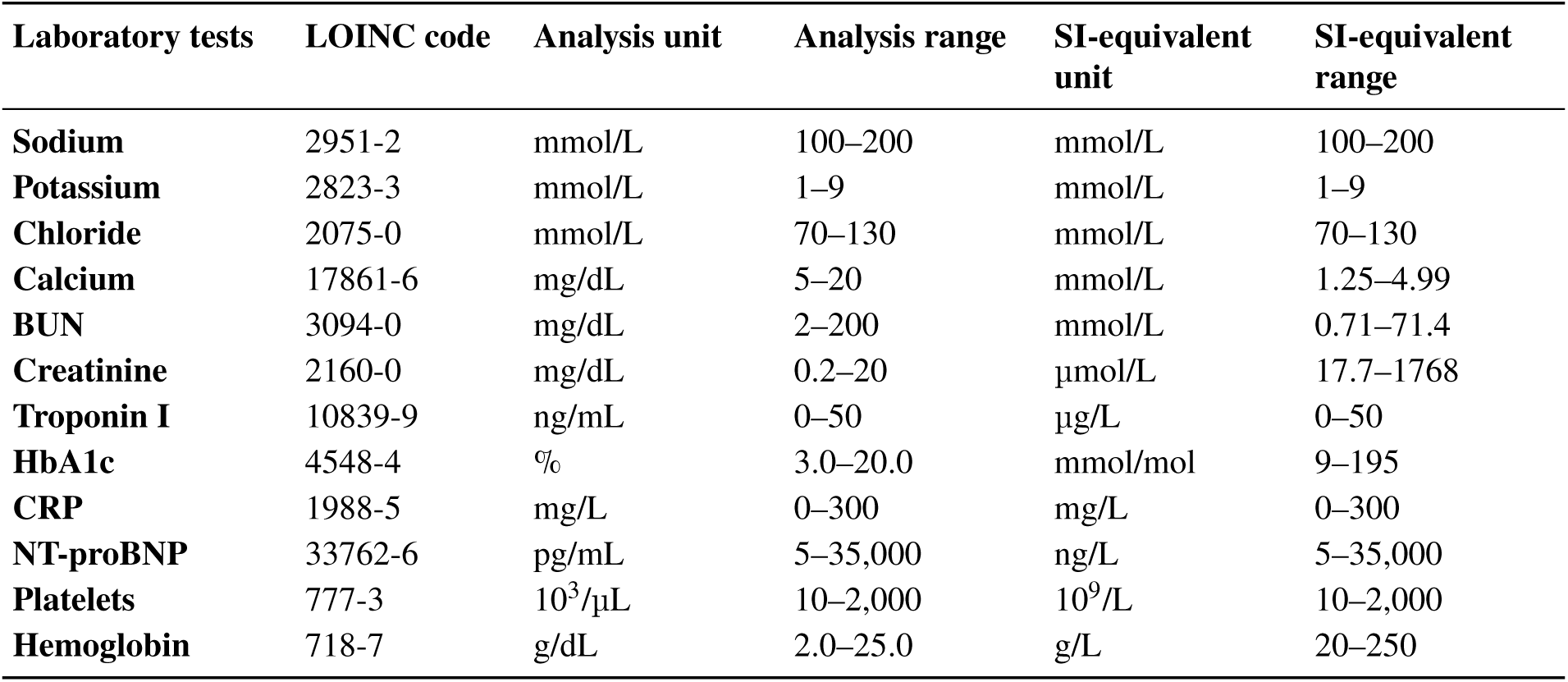
LOINC codes, original units, SI-equivalent units and biologically plausible ranges for laboratory tests. Laboratory tests were identified using LOINC codes available in the source electronic health record data. The Unit and Range columns reflect the standardized units and biologically plausible ranges used during data preprocessing. Corresponding SI-equivalent units and ranges are provided for reporting purposes. Values outside the specified biologically plausible ranges were treated as implausible and excluded prior to imputation. Ranges were defined based on clinical plausibility rather than population-specific reference intervals. SI, International System of Units; BUN, blood urea nitrogen; CRP, C-reactive protein; HbA1c, hemoglobin A1c; LOINC, Logical Observation Identifiers Names and Codes; NT-proBNP, N-terminal pro-B-type natriuretic peptide.

| Laboratory tests | LOINC code | Analysis unit | Analysis range | SI-equivalent unit | SI-equivalent range |
|---|---|---|---|---|---|
| **Sodium** | 2951-2 | mmol/L | 100–200 | mmol/L | 100–200 |
| **Potassium** | 2823-3 | mmol/L | 1–9 | mmol/L | 1–9 |
| **Chloride** | 2075-0 | mmol/L | 70–130 | mmol/L | 70–130 |
| **Calcium** | 17861-6 | mg/dL | 5–20 | mmol/L | 1.25–4.99 |
| **BUN** | 3094-0 | mg/dL | 2–200 | mmol/L | 0.71–71.4 |
| **Creatinine** | 2160-0 | mg/dL | 0.2–20 | µmol/L | 17.7–1768 |
| **Troponin I** | 10839-9 | ng/mL | 0–50 | µg/L | 0–50 |
| **HbA1c** | 4548-4 | % | 3.0–20.0 | mmol/mol | 9–195 |
| **CRP** | 1988-5 | mg/L | 0–300 | mg/L | 0–300 |
| **NT-proBNP** | 33762-6 | pg/mL | 5–35,000 | ng/L | 5–35,000 |
| **Platelets** | 777-3 | 10<sup>3</sup>/µL | 10–2,000 | 10<sup>9</sup>/L | 10–2,000 |
| **Hemoglobin** | 718-7 | g/dL | 2.0–25.0 | g/L | 20–250 |

**Supplementary Table S4.** Units and biologically plausible ranges for vital sign and anthropometric measurements. Values outside the specified biologically plausible ranges were treated as implausible and excluded prior to imputation. Ranges were defined based on clinical plausibility rather than population-specific reference intervals. BMI, body mass index.

| **Feature** | **Unit** | **Range** |
|---|---|---|
| **Pulse rate** | beats/min | 20–250 |
| **Respiration rate** | breaths/min | 4–90 |
| **Systolic blood pressure** | mm Hg | 50–300 |
| **Diastolic blood pressure** | mm Hg | 30–200 |
| **BMI** | kg/m<sup>2</sup> | 7–90 |

**Supplementary Table S5.** Distribution and missingness of the 30 SHAP-selected features used in the final XGBoost model by arrhythmia status. Continuous variables are reported as median (IQR) among patients with available measurements and in original data units, rather than converted to SI units; binary variables are reported as n (%). Missingness was assessed before median imputation of continuous measurement-derived features. Normalized delta features represent change per day; 18-month normalized delta features were calculated over a fixed observation window. BMI, body mass index; IQR, interquartile range; SHAP, SHapley Additive exPlanations.

| **Feature** | **Overall cohort<br>(n=30,356)** | **Arrhythmia-negative<br>(n=23,521)** | **Arrhythmia-positive<br>(n=6,835)** | **Missing overall, n (%)** |
|---|---|---|---|---|
| **Age (years)** | 72.0<br>(64.0–79.0) | 70.0<br>(60.0–78.0) | 78.0<br>(72.0–84.0) | 0 (0.0) |
| **Platelets, median (10<sup>3</sup>/µL)** | 246.0<br>(207.0–291.0) | 247.0<br>(208.0–292.0) | 236.0<br>(196.0–283.0) | 7,509 (24.7) |
| **Hemoglobin, normalized delta (g/dL)** | -0.0001<br>(-0.0009–0.0004) | -0.0001<br>(-0.0009–0.0004) | 0.0000<br>(-0.0012–0.0004) | 7,330 (24.1) |
| **Blood urea nitrogen, normalized delta (mg/dL)** | 0.0000<br>(-0.0020–0.0038) | 0.0000<br>(-0.0019–0.0037) | 0.0000<br>(-0.0026–0.0054) | 7,285 (24.0) |
| **Other racial and ethnic groups** | 19,525 (64.3) | 16,027 (68.1) | 3,498 (51.2) | 0 (0.0) |
| **Platelets, normalized delta (10<sup>3</sup>/µL)** | 0.0000<br>(-0.0262–0.0170) | 0.0000<br>(-0.0256–0.0169) | 0.0000<br>(-0.0323–0.0180) | 7,509 (24.7) |
| **Blood urea nitrogen, median (mg/dL)** | 16.0<br>(13.0–19.0) | 15.5<br>(13.0–19.0) | 17.0<br>(13.5–21.0) | 7,285 (24.0) |
| **Hemoglobin, median (g/dL)** | 13.9<br>(12.9–15.0) | 14.0<br>(13.0–15.0) | 13.9<br>(12.8–15.0) | 7,330 (24.1) |
| **Calcium, normalized delta (mg/dL)** | 0.0000<br>(-0.0003–0.0002) | 0.0000<br>(-0.0003–0.0002) | 0.0000<br>(-0.0003–0.0003) | 7,305 (24.1) |
| **Chloride, normalized delta (mmol/L)** | 0.0000<br>(-0.0017–0.0021) | 0.0000<br>(-0.0017–0.0021) | 0.0000<br>(-0.0024–0.0028) | 7,300 (24.0) |
| **Respiration rate, median (breaths/min)** | 16.0<br>(16.0–17.0) | 16.0<br>(16.0–17.0) | 16.0<br>(16.0–17.0) | 3,611 (11.9) |
| **Potassium, normalized delta (mmol/L)** | 0.0000<br>(-0.0002–0.0003) | 0.0000<br>(-0.0002–0.0003) | 0.0000<br>(-0.0003–0.0004) | 7,299 (24.0) |
| **Creatinine, 18-month normalized delta (mg/dL)** | 0.0000<br>(-0.0002–0.0002) | 0.0000<br>(-0.0002–0.0002) | 0.0000<br>(-0.0002–0.0002) | 5,351 (17.6) |
| **Sodium, normalized delta (mmol/L)** | 0.0000<br>(-0.0017–0.0017) | 0.0000<br>(-0.0017–0.0017) | 0.0000<br>(-0.0022–0.0024) | 7,299 (24.0) |
| **Hemoglobin, median of 3 most recent measurements (g/dL)** | 13.9<br>(12.9–15.0) | 13.9<br>(12.9–15.0) | 13.9<br>(12.7–15.0) | 7,330 (24.1) |
| **BMI, mean of 3 most recent measurements (kg/m<sup>2</sup>)** | 28.33<br>(24.74–32.57) | 28.31<br>(24.66–32.52) | 28.38<br>(25.06–32.78) | 3,611 (11.9) |
| **Calcium, mean of 3 most recent measurements (mg/dL)** | 9.5<br>(9.3–9.8) | 9.5<br>(9.3–9.8) | 9.5<br>(9.2–9.8) | 7,305 (24.1) |
| **Pulse rate, normalized delta (beats/min)** | 0.0000<br>(-0.0050–0.0045) | 0.0000<br>(-0.0047–0.0041) | 0.0000<br>(-0.0071–0.0074) | 3,611 (11.9) |
| **Congestive heart failure** | 1,854 (6.1) | 1,343 (5.7) | 511 (7.5) | 0 (0.0) |
| **Blood urea nitrogen, mean of 3 most recent measurements (mg/dL)** | 16.0<br>(13.0–20.0) | 15.7<br>(12.7–19.7) | 17.0<br>(13.5–21.0) | 7,285 (24.0) |
| **Pulse rate, median (beats/min)** | 76.0<br>(69.5–83.0) | 76.0<br>(70.0–83.0) | 74.0<br>(68.0–82.0) | 3,611 (11.9) |
| **Hemoglobin, 18-month normalized delta (g/dL)** | 0.0000<br>(-0.0014–0.0010) | 0.0000<br>(-0.0014–0.0010) | 0.0000<br>(-0.0017–0.0007) | 7,330 (24.1) |
| **Female** | 15,474 (51.0) | 12,340 (52.5) | 3,134 (45.9) | 0 (0.0) |
| **Chloride, median of 3 most recent measurements (mmol/L)** | 102.0<br>(100.0–104.0) | 102.0<br>(100.0–104.0) | 102.0<br>(100.0–104.0) | 7,300 (24.0) |
| **BMI, median of 3 most recent measurements (kg/m<sup>2</sup>)** | 28.34<br>(24.74–32.59) | 28.32<br>(24.66–32.55) | 28.40<br>(25.06–32.81) | 3,611 (11.9) |
| **White, non-Hispanic** | 10,831 (35.7) | 7,494 (31.9) | 3,337 (48.8) | 0 (0.0) |
| **Calcium, median of 3 most recent measurements (mg/dL)** | 9.5<br>(9.2–9.8) | 9.5<br>(9.2–9.8) | 9.5<br>(9.2–9.8) | 7,305 (24.1) |
| **BMI, normalized delta (kg/m<sup>2</sup>)** | 0.0000<br>(-0.0009–0.0004) | 0.0000<br>(-0.0009–0.0004) | 0.0000<br>(-0.0013–0.0006) | 3,611 (11.9) |
| **BMI, median (kg/m<sup>2</sup>)** | 28.48<br>(25.00–32.77) | 28.48<br>(24.95–32.76) | 28.48<br>(25.27–32.79) | 3,611 (11.9) |
| **Respiration rate, median of 3 most recent measurements (breaths/min)** | 16.0<br>(16.0–17.0) | 16.0<br>(16.0–17.5) | 16.0<br>(16.0–17.0) | 3,611 (11.9) |

**Supplementary Table S6.**
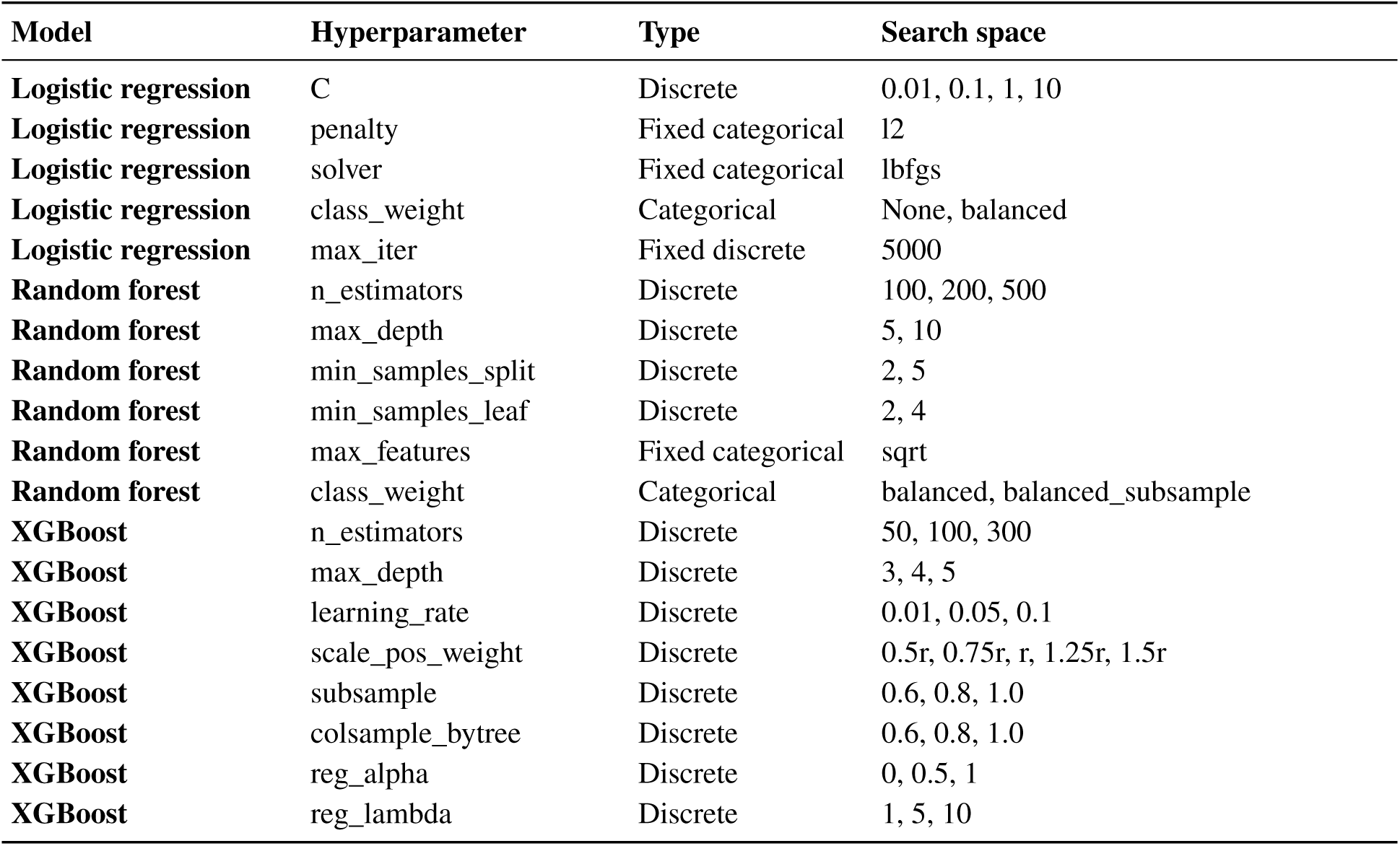
Hyperparameter search space used for model tuning. Hyperparameter tuning was performed using an exhaustive grid search with a 3-fold stratified cross-validation on the training set. AUROC was used as the optimization metric. For XGBoost, r denotes the observed ratio of arrhythmia-negative to arrhythmia-positive patients in the training set. This ratio was 16,943 / 4,901 = 3.46. The evaluated scale_pos_weight values corresponded to 0.5, 0.75, 1.0, 1.25, and 1.5 times r. Fixed parameters were held constant during grid search. AUROC, area under the receiver operating characteristic curve.

**Supplementary Table S7.** Characterization of the clinical-score benchmark cohort. The common score-calculable subset consisted of holdout patients for whom both CHARGE-AF and HARMS_2_-AF could be calculated. The final I48 benchmark cohort was obtained by retaining I48-positive patients and original arrhythmia-negative controls and excluding 289 patients positive for non-I48 arrhythmias. Overall, 89.3% of the holdout cohort was retained in the common calculable subset. Retention was 95.1% among original negatives, 70.1% among all arrhythmia-positive patients, and 59.3% among I48-positive patients. IQR, interquartile range; SD, standard deviation; CHARGE-AF, Cohorts for Heart and Aging in Genomic Epidemiology. *In the final benchmark cohort, all retained arrhythmia-positive patients were I48-positive by design.

| Characteristic | Full holdout (n=3,051) | Common score-calculable subset (n=2,725) | Excluded due to score calculability (n=326) | Final I48 benchmark cohort (n=2,436) |
|---|---|---|---|---|
| **Age, mean (SD), years** | 69.8 (15.4) | 69.6 (15.0) | 71.3 (18.0) | 68.9 (15.3) |
| **Age, median (IQR), years** | 73 (64–80) | 72 (64–79) | 76 (65–84) | 72 (63–79) |
| **Female, n (%)** | 1,565 (51.3) | 1,397 (51.3) | 168 (51.5) | 1,269 (52.1) |
| **Male, n (%)** | 1,486 (48.7) | 1,328 (48.7) | 158 (48.5) | 1,167 (47.9) |
| **White, non-Hispanic, n (%)** | 1,121 (36.7) | 1,003 (36.8) | 118 (36.2) | 859 (35.3) |
| **Other racial and ethnic groups, n (%)** | 1,930 (63.3) | 1,722 (63.2) | 208 (63.8) | 1,577 (64.7) |
| **Composite arrhythmia-positive, n (%)** | 709 (23.2) | 497 (18.2) | 212 (65.0) | 208 (8.5) * |
| **I48-positive, n (%)** | 351 (11.5) | 208 (7.6) | 143 (43.9) | 208 (8.5) |
| **Original arrhythmia-negative, n (%)** | 2,342 (76.8) | 2,228 (81.8) | 114 (35.0) | 2,228 (91.5) |

**Supplementary Table S8.** Lead-time sensitivity analysis of final XGBoost model performance on the primary holdout test set. The same 3,051 held-out patients were evaluated at each lead time. Features were regenerated using clinical information available only up to the specified number of days before the index date, and the previously trained final XGBoost model was applied without retraining. Sensitivity, specificity, precision, NPV, F1 score, and accuracy were calculated using the default probability threshold of 0.5. AUROC and AUPRC are threshold-independent. All performance metrics are reported with 95% confidence intervals. CI, confidence interval; AUROC, area under the receiver operating characteristic curve; AUPRC, area under the precision-recall curve; NPV, negative predictive value.

| Metric | 1 day | 30 days | 90 days | 180 days |
|---|---|---|---|---|
| **N evaluated** | 3,051 | 3,051 | 3,051 | 3,051 |
| **Arrhythmia-positive, n (%)** | 709 (23.2) | 709 (23.2) | 709 (23.2) | 709 (23.2) |
| **AUROC (95% CI)** | 0.8935<br>(0.8786–0.9077) | 0.8968<br>(0.8822–0.9112) | 0.8963<br>(0.8818–0.9104) | 0.8963<br>(0.8816–0.9098) |
| **AUPRC (95% CI)** | 0.7922<br>(0.7626–0.8215) | 0.7940<br>(0.7686–0.8208) | 0.7936<br>(0.7679–0.8203) | 0.7840<br>(0.7578–0.8120) |
| **Sensitivity (95% CI)** | 0.5642<br>(0.5274–0.6002) | 0.5910<br>(0.5544–0.6266) | 0.6164<br>(0.5800–0.6514) | 0.6432<br>(0.6072–0.6776) |
| **Specificity (95% CI)** | 0.9714<br>(0.9638–0.9774) | 0.9637<br>(0.9553–0.9706) | 0.9577<br>(0.9488–0.9652) | 0.9441<br>(0.9340–0.9527) |
| **Precision (95% CI)** | 0.8565<br>(0.8218–0.8854) | 0.8313<br>(0.7962–0.8615) | 0.8153<br>(0.7802–0.8459) | 0.7768<br>(0.7414–0.8087) |
| **Negative predictive value (95% CI)** | 0.8804<br>(0.8673– 0.8924) | 0.8861<br>(0.8732– 0.8979) | 0.8918<br>(0.8791– 0.9034) | 0.8973<br>(0.8847– 0.9087) |
| **F1 score (95% CI)** | 0.6803<br>(0.648– 0.7121) | 0.6908<br>(0.6601– 0.7201) | 0.7020<br>(0.6743– 0.7296) | 0.7037<br>(0.6756– 0.732) |
| **Accuracy (95% CI)** | 0.8768<br>(0.8646– 0.888) | 0.8771<br>(0.865– 0.8883) | 0.8784<br>(0.8663– 0.8895) | 0.8741<br>(0.8619– 0.8854) |
| **Brier score (95% CI)** | 0.0989<br>(0.0924– 0.105) | 0.098<br>(0.0917– 0.104) | 0.0981<br>(0.0922– 0.1039) | 0.1000<br>(0.0938– 0.1058) |

**Supplementary Table S9.** Performance of the final XGBoost model by arrhythmia target group on the primary holdout test set. Each target-group analysis treated patients with the specified arrhythmia group as positive and patients from the original arrhythmia-negative holdout population as controls. Patients positive for other arrhythmia groups were excluded from that comparison. The same 2,342 arrhythmia-negative controls were used across all target-group analyses. Sensitivity, specificity, precision, NPV, F1 score, and accuracy were calculated using the default probability threshold of 0.5. AUROC and AUPRC are threshold-independent. All performance metrics are reported with 95% confidence intervals. CI, confidence interval; AUROC, area under the receiver operating characteristic curve; AUPRC, area under the precision-recall curve; NPV, negative predictive value.

| **Measure** | **I44** | **I47** | **I48** | **I49** |
|---|---|---|---|---|
| **N evaluated** | 2,410 | 2,493 | 2,693 | 2,481 |
| **Arrhythmia-positive, n** | 68 | 151 | 351 | 139 |
| **Arrhythmia -negative, n** | 2,342 | 2,342 | 2,342 | 2,342 |
| **AUROC (95% CI)** | 0.8885 (0.8452–0.9318) | 0.8336 (0.7964–0.8708) | 0.9218 (0.9047–0.9389) | 0.8893 (0.8590–0.9196) |
| **AUPRC (95% CI)** | 0.4119 (0.3047–0.5536) | 0.4283 (0.3518–0.5194) | 0.7656 (0.7200–0.8083) | 0.5378 (0.4473–0.6312) |
| **Sensitivity (95% CI)** | 0.5588 (0.4408–0.6705) | 0.4238 (0.3479–0.5036) | 0.6382 (0.5867–0.6867) | 0.5324 (0.4497–0.6133) |
| **Specificity (95% CI)** | 0.9714 (0.9638–0.9774) | 0.9714 (0.9638–0.9774) | 0.9714 (0.9638–0.9774) | 0.9714 (0.9638–0.9774) |
| **Precision (95% CI)** | 0.3619 (0.2764–0.4572) | 0.4885 (0.4045–0.5732) | 0.7698 (0.7181–0.8144) | 0.5248 (0.4428–0.6055) |
| **NPV (95% CI)** | 0.9870 (0.9815–0.9909) | 0.9632 (0.9548–0.9700) | 0.9471 (0.9374–0.9554) | 0.9722 (0.9648–0.9781) |
| **F1 score (95% CI)** | 0.4393 (0.3508–0.5283) | 0.4539 (0.3813–0.5227) | 0.6978 (0.6590–0.7367) | 0.5286 (0.4542–0.6035) |
| **Accuracy (95% CI)** | 0.9598 (0.9511–0.9669) | 0.9382 (0.9281–0.9470) | 0.9280 (0.9176–0.9371) | 0.9468 (0.9373–0.9550) |
| **Brier score (95% CI)** | 0.0473 (0.0436–0.0510) | 0.0611 (0.0559–0.0667) | 0.0663 (0.0617–0.0711) | 0.0550 (0.0508–0.0597) |

**Supplementary Table S10.**
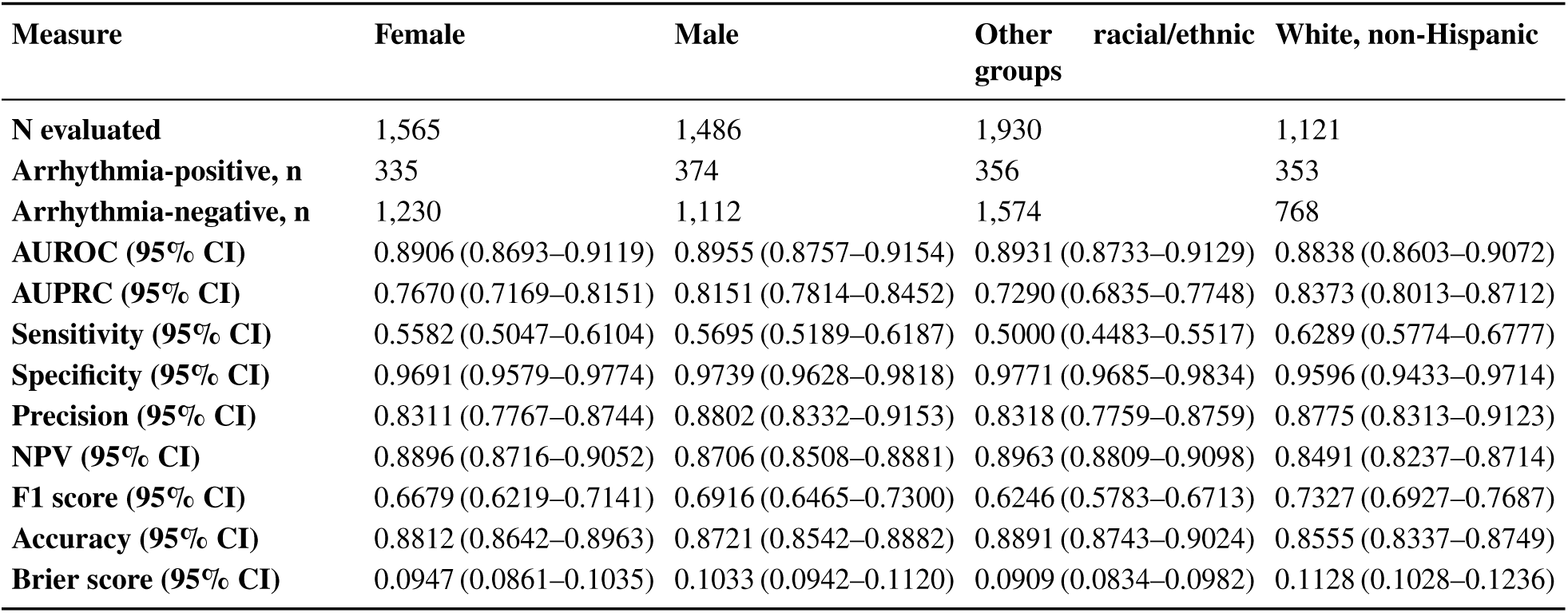
XGBoost performance across gender and race/ethnicity groups on the primary holdout test set. Performance of the XGBoost model stratified by gender and race/ethnicity subgroups. Sensitivity, specificity, precision, NPV, F1 score, and accuracy were calculated using the default probability threshold of 0.5. AUROC and AUPRC are threshold-independent. All performance metrics are reported with 95% confidence intervals. CI, confidence interval; AUROC, area under the receiver operating characteristic curve; AUPRC, area under the precision-recall curve; NPV, negative predictive value.

| Measure | Female | Male | Other racial/ethnic groups | White, non-Hispanic |
|---|---|---|---|---|
| **N evaluated** | 1,565 | 1,486 | 1,930 | 1,121 |
| **Arrhythmia-positive, n** | 335 | 374 | 356 | 353 |
| **Arrhythmia-negative, n** | 1,230 | 1,112 | 1,574 | 768 |
| **AUROC (95% CI)** | 0.8906 (0.8693–0.9119) | 0.8955 (0.8757–0.9154) | 0.8931 (0.8733–0.9129) | 0.8838 (0.8603–0.9072) |
| **AUPRC (95% CI)** | 0.7670 (0.7169–0.8151) | 0.8151 (0.7814–0.8452) | 0.7290 (0.6835–0.7748) | 0.8373 (0.8013–0.8712) |
| **Sensitivity (95% CI)** | 0.5582 (0.5047–0.6104) | 0.5695 (0.5189–0.6187) | 0.5000 (0.4483–0.5517) | 0.6289 (0.5774–0.6777) |
| **Specificity (95% CI)** | 0.9691 (0.9579–0.9774) | 0.9739 (0.9628–0.9818) | 0.9771 (0.9685–0.9834) | 0.9596 (0.9433–0.9714) |
| **Precision (95% CI)** | 0.8311 (0.7767–0.8744) | 0.8802 (0.8332–0.9153) | 0.8318 (0.7759–0.8759) | 0.8775 (0.8313–0.9123) |
| **NPV (95% CI)** | 0.8896 (0.8716–0.9052) | 0.8706 (0.8508–0.8881) | 0.8963 (0.8809–0.9098) | 0.8491 (0.8237–0.8714) |
| **F1 score (95% CI)** | 0.6679 (0.6219–0.7141) | 0.6916 (0.6465–0.7300) | 0.6246 (0.5783–0.6713) | 0.7327 (0.6927–0.7687) |
| **Accuracy (95% CI)** | 0.8812 (0.8642–0.8963) | 0.8721 (0.8542–0.8882) | 0.8891 (0.8743–0.9024) | 0.8555 (0.8337–0.8749) |
| **Brier score (95% CI)** | 0.0947 (0.0861–0.1035) | 0.1033 (0.0942–0.1120) | 0.0909 (0.0834–0.0982) | 0.1128 (0.1028–0.1236) |

**Supplementary Table S11.** Age-stratified performance of the final XGBoost model on the primary holdout test set. Performance of the XGBoost model across prespecified patient age groups, including subgroup sample sizes and arrhythmia case distribution. Precision is not reported for the 18–39 age group because there were no predicted positive cases at the default threshold. Sensitivity, specificity, precision, NPV, F1 score, and accuracy were calculated using the default probability threshold of 0.5. AUROC and AUPRC are threshold-independent. All performance metrics are reported with 95% confidence intervals. CI, confidence interval; AUROC, area under the receiver operating characteristic curve; AUPRC, area under the precision-recall curve; NPV, negative predictive value.

| **Measure** | **18–39 years** | **40–64 years** | **65–79 years** | **≥80 years** |
|---|---|---|---|---|
| **N evaluated** | 200 | 641 | 1,495 | 715 |
| **Arrhythmia-positive, n** | 5 | 61 | 354 | 289 |
| **Arrhythmia-negative, n** | 195 | 580 | 1,141 | 426 |
| **AUROC (95% CI)** | 0.5564 (0.2819–0.8309) | 0.8442 (0.7882–0.9002) | 0.8694 (0.8463–0.8925) | 0.9019 (0.8771–0.9267) |
| **AUPRC (95% CI)** | 0.0914 (0.0105–0.5000) | 0.4882 (0.3655–0.6100) | 0.7614 (0.7209–0.8021) | 0.8807 (0.8412–0.9151) |
| **Sensitivity (95% CI)** | 0.0000 (0.0000–0.4345) | 0.1967 (0.1163–0.3131) | 0.5056 (0.4538–0.5574) | 0.7232 (0.6689–0.7716) |
| **Specificity (95% CI)** | 1.0000 (0.9807–1.0000) | 0.9914 (0.9800–0.9963) | 0.9684 (0.9566–0.9771) | 0.9390 (0.9121–0.9580) |
| **Precision (95% CI)** | NA | 0.7059 (0.4687–0.8672) | 0.8326 (0.7769–0.8765) | 0.8894 (0.8428–0.9234) |
| **NPV (95% CI)** | 0.9750 (0.9428–0.9893) | 0.9215 (0.8977–0.9401) | 0.8633 (0.8434–0.8810) | 0.8333 (0.7974–0.8640) |
| **F1 score (95% CI)** | 0.0000 (0.0000–0.0000) | 0.3077 (0.1639–0.4272) | 0.6292 (0.5791–0.6752) | 0.7977 (0.7567–0.8316) |
| **Accuracy (95% CI)** | 0.9750 (0.9428–0.9893) | 0.9158 (0.8917–0.9349) | 0.8589 (0.8403–0.8756) | 0.8517 (0.8238–0.8759) |
| **Brier score (95% CI)** | 0.0255 (0.0081–0.0447) | 0.0675 (0.0541–0.0810) | 0.1118 (0.1026–0.1215) | 0.1206 (0.1083–0.1334) |

**Supplementary Figure S1.**
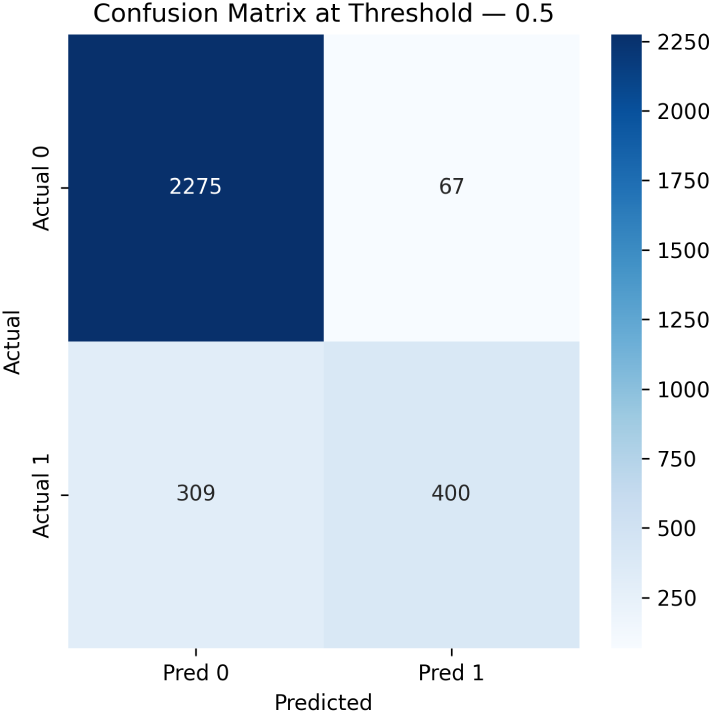
Confusion matrix for the final XGBoost model on the holdout test set at a classification threshold of 0.5. Actual 0: arrhythmia-negative; Actual 1: arrhythmia-positive; Pred 0: predicted negative; Pred 1: predicted positive.

**Supplementary Figure S2.**
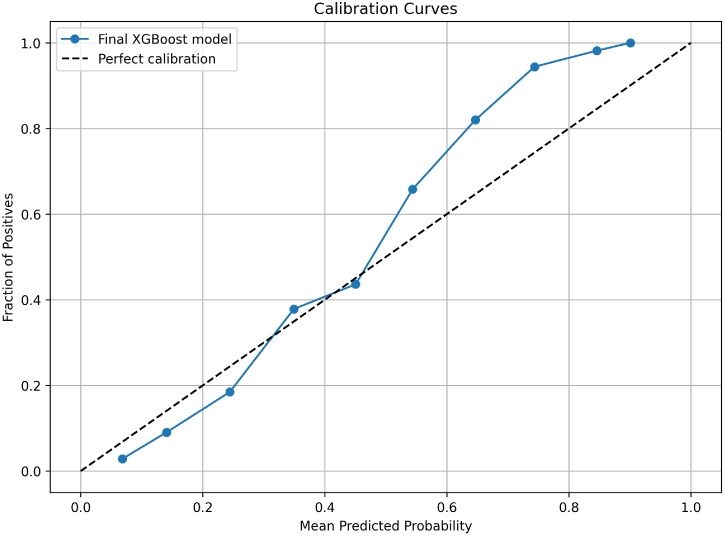
Calibration curve on the holdout test set.

**Supplementary Figure S3.**
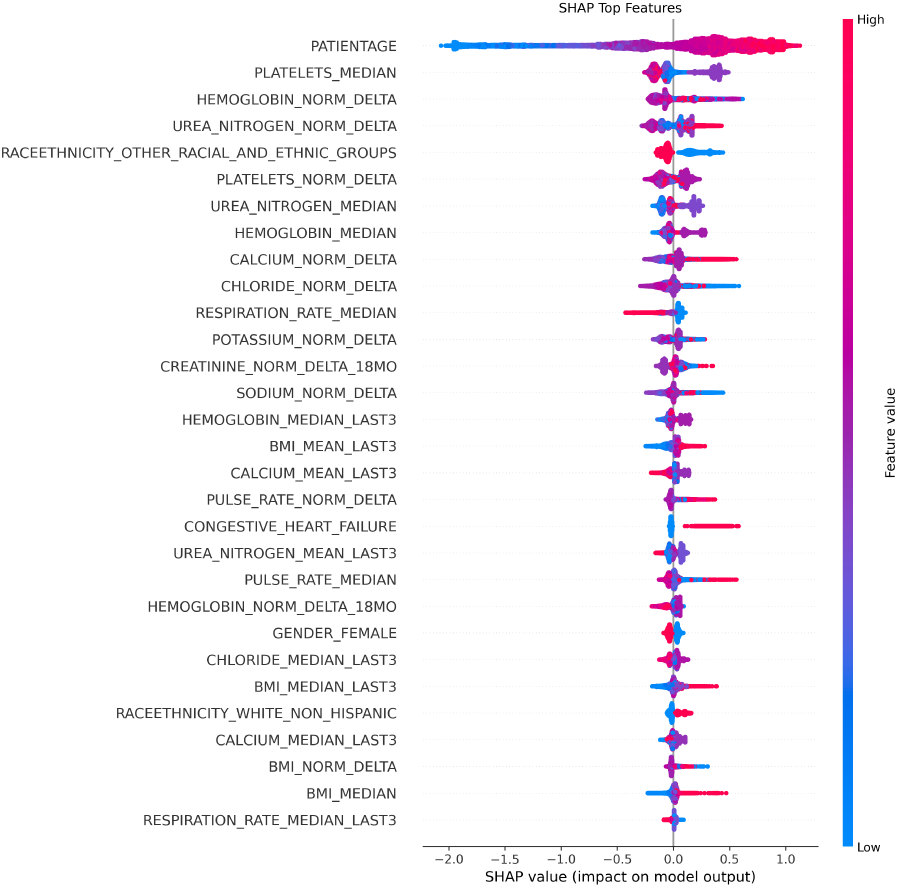
Shapley additive explanations beeswarm plot for all 30 SHAP-selected features in the final XGBoost model, ordered by mean absolute SHAP value. Each point represents 1 patient; horizontal position indicates the direction and magnitude of that feature’s contribution to the model output, and the color scale encodes the underlying feature value from low to high. SHAP: Shapley additive explanations; XGBoost: extreme gradient boosting.

